# Evaluation of Real-World Mobility Recovery after Hip Fracture using Digital Mobility Outcomes

**DOI:** 10.1101/2024.05.31.24308265

**Authors:** Monika Engdal, Kristin Taraldsen, Carl-Philipp Jansen, Raphael Simon Peter, Beatrix Vereijken, Clemens Becker, Jorunn L Helbostad, Jochen Klenk

**Affiliations:** Department of Neuromedicine and Movement Science, NTNU, Trondheim, Norway; Department of Rehabilitation Science and Health Technology, OsloMet, Oslo, Norway; Department of Clinical Gerontology, Robert Bosch Hospital, Stuttgart, Germany; Geriatric Department, Heidelberg University Medical Center, Heidelberg, Germany; Institute of Epidemiology and Medical Biometry, Ulm University, Ulm, Germany; IB University of Health and Social Sciences, Study Center Stuttgart, Stuttgart, Germany

## Abstract

**Background:** The main focus of rehabilitation following hip fracture is to regain mobility.

**Objectives:** To estimate the progression of real-world mobility the first year after hip fracture using digital mobility outcomes.

**Design:** An exploratory, prospective cohort study with pooled data from four previously conducted clinical trials.

**Setting and Subjects:** We combined data from the Trondheim Hip Fracture Trial and Eva-Hip Trial in Trondheim, Norway, and the PROFinD 1 and PROFinD 2 trials in Stuttgart and Heidelberg, Germany, resulting in a sample of 717 hip fracture patients aged ≥65 years.

**Methods:** Each of the trials assessed mobility using body-fixed sensors (activPAL™) at three time points, collectively providing observations across the entire first year post-surgery. The following 24-hour DMOs were calculated: total walking duration (minutes), maximum number of steps within a walking bout, and number of sit-to-stand-to-walk transfers. Continuous one-year progression of the median, the 25^th^ percentile, and the 75^th^ percentile were estimated using quantile regression models with splines.

**Results:** The dataset contained 5,909 observation days. The median daily total walking duration increased until 36 weeks post-surgery reaching 40 minutes; daily maximum number of steps within a walking bout increased during the first eight weeks and then stabilized at less than 100 steps; daily sit-to-stand-to-walk transfers reached a plateau after six weeks with less than 40 transfers.

**Conclusions:** The three DMOs progressed differently and attained plateau levels at varying times during the first year after hip fracture, indicating that these Digital Mobility Outcomes provide complementary information about different aspects of mobility recovery.

## Introduction

Each year, 1.6 million people worldwide suffer from hip fractures [1], with a projected estimate of 6 million hip fractures annually by 2050 [1, 2]. A hip fracture has severe consequences for older adults, leading to short- and long-term mobility disability and related limitations in daily functioning [3, 4]. A review from 2016 found that only 40-60% of hip fracture survivors recover to their pre-fracture level of mobility and ability to perform activities of daily living. Additionally, 10-20% of survivors require long-term nursing home care 6-12 months post-surgery [5]. Mobility is a crucial aspect of human life and a significant marker of health and function [6, 7]. Therefore, the main focus of rehabilitation following hip fracture is to regain mobility and sustain the ability to carry out activities of daily living to ensure independent living [8].

Traditionally, mobility has been assessed using patient-reported outcomes through questionnaires or physical performance assessments in laboratory or clinical settings. However, body-worn sensors now accurately measure mobility parameters in real-world environments, providing continuous data on walking and other digital mobility outcomes (DMOs) [9, 10]. While self-reports can be biased by inaccurate recall and performance-based tests only provide brief snapshots of patients’ capability, real-world mobility measures capture high-granularity information on what patients are actually doing in daily life over longer periods of time. Thus, DMOs can provide valuable information about mobility recovery after hip fracture.

Despite mobility being a major challenge and an important focus of rehabilitation after a hip fracture [11, 12], a recent review found that most interventions are not designed to evaluate effects on real-world mobility [13]. The review found gait speed from short walks in a laboratory setting to be the most commonly used parameter, and often the only mobility outcome in hip fracture trials. Only four of 29 included trials reported real-world DMOs [14–18]. One exercise study found significant effects on the primary outcome gait speed, but no transfer effects to real-world upright time [14]. Similarly, an observational study on hip fracture patients attending geriatric rehabilitation, found that in-lab physical capacity tests, including gait speed, only had a fair to modest correlation with real-world DMOs [19]. To gain deeper insight into recovery of mobility in daily life, mobility needs to be explored beyond in-lab capacity tests. Therefore, the objective of this study was to estimate the progression of real-world mobility using multiple DMOs during the first year after hip fracture. To provide information on gait volume, walking performance, and transfers between activities, we chose the following 24-hour DMOs: total walking duration, maximum number of continuous steps within a walking bout, and number of sit-to-stand-to-walk transfers.

## Methods

### Design

To address the challenge of limited longitudinal data availability, the present study adopted a prospective cohort design with pooled data from four previously conducted randomized controlled trials: The Trondheim Hip Fracture Trial [15, 20] and the Eva-Hip Trial [14] in Trondheim, Norway, along with the PROFinD 1 [21] and PROFinD 2 [22] trials in Stuttgart and Heidelberg, Germany. This approach fascilitated secondary, exploratory data analyses of real-world mobility among hip fracture patients during the first year after surgery. The intervention in all trials focused on improving mobility and activity in daily life, and all control groups underwent rehabilitation as usual.

### Setting and Sample

The Trondheim Hip Fracture Trial was conducted to examine whether comprehensive geriatric care in a specialized orthogeriatric unit during hospital stay improved recovery and was more cost-effective than standard orthopedic unit treatment for hip fracture patients [23]. The study included 397 community-dwelling older adults (≥70 years) from April 2008 to December 2010. The Eva-Hip Trial aimed to evaluate the clinical effectiveness and cost-effectiveness of functional exercise 4-6 months after hip fracture surgery, compared to practice as usual [24]. The study included 143 community-dwelling older adults (≥70 years) from February 2011 until March 2014. The PROFinD 1 Trial was conducted to investigate whether step-by-step in-patient rehabilitation 3-8 weeks post-surgery could increase physical activity and fall-related self-efficacy in hip- and pelvic fracture patients with fear of falling, compared to standard in-patient rehabilitation [25]. The study included 111 community-dwelling adults (≥ 60 years) from April 2011 until December 2013. The PROFinD 2 Trial involved 185 community-dwelling hip- and pelvic fracture patients (≥ 65 years) with cognitive impairment according to Mini-Mental State Examination (MMSE; scores between 17-26). The study aimed to compare the effect of a four month post-discharge multifactorial home-based rehabilitation program 4-7 months post-surgery on physical activity and functional performance with usual care and was conducted between July 2015 and February 2018 [22]. Only the hip fracture patients in the PROFinD 1 and PROFinD 2 trials were included in the current analyses.

### Descriptive measures

Demographics and clinical characteristics included age, gender, Body Mass Index (BMI), living alone at admission, indoor falls, type of fracture, and preferred gait speed from the Short Physical Performance Battery’s (SPPB) 4-meter walk at four months (six months for the PROFinD 2 Trial) post-surgery [26]. Cognitive function was assessed by Mini-Mental State Examination (MMSE) [27] or Short Orientation Memory Concentration test (SOMC) [28], at four months post-surgery for the Trondheim cohorts and the third week post-surgery for the Stuttgart cohorts.

### Digital Mobility Outcomes (DMOs)

Real-world mobility was measured using body-fixed accelerometer-based sensors (activPAL™, PAL Technologies Ltd., Glasgow, UK). The device was attached to the non-affected front thigh using waterproof tape, and worn continuously for a minimum of 24 hours. The activPAL’s software default settings were used to program the sensors and to process the recorded data, i.e., upright events were established with a minimum length of 10 sec. The software algorithms categorize accelerometer data into three activities: (1) sitting/ lying, (2) standing, and (3) walking. Based on this event-based data output, we calculated the following DMOs for each valid day (i.e., consisting of 24 hours of recording):

Total walking duration (minutes), number of sit-to-stand-to-walk transfers, and the maximum number of continuous steps within a walking bout, as derived by the activPAL software. A previous validation study with hip fracture patients found high accuracy (100%) in classifying activities and recognizing sit-to-stand transfers, but underestimation of step counts and walking duration at slow gait speeds (≤0.47 m/sec) [29].

All trials monitored mobility at three time points, collectively providing observations across the entire first year post-surgery. In the Trondheim Hip Fracture Trial, DMOs from the hospital setting the fourth postoperative day and from four-day assessments at four and 12 months post-surgery were collected [23]. In the Eva-Hip Trial, four-day assessments of DMOs at four, six, and 12 months post-surgery were collected [24]. The PROFinD 1 Trial collected one-day recordings at weeks 2-3 and again six weeks post-surgery during an in-patient rehabilitation stay, and then for seven days at 4-5 months post-surgery [25]. In the PROFinD 2 Trial, DMOs from three days at 2-3 months, 6-7 months, and 10-11 months post-surgery were collected [22].

### Data Analysis and Statistics

To verify each 24-hour recording, visual inspection of the DMOs was done by two authors, and all days with full 24-hour recordings were included in the dataset. The number of valid days for each participant and at each time point varied between one and seven consecutive days. The DMOs at all assessment time points across the trials were merged into one database, providing a longitudinal dataset covering the entire one year observation period. The R 4.2.2. statistical package (with package quantregGrowth 1.7.0) was used to analyse mobility data and the SPSS Statistics 28.0 (IBM, NY, USA) for descriptive analyses. Continuous variables were summarised as means and standard deviations (SDs) or median and interquartile range (IQR), and categorical variables were presented as frequencies and proportions.

In the first step, we estimated the continuous progression of the 25^th,^ 50^th^ (median), and 75^th^ percentiles using quantile regression models with splines for each considered DMO within the entire population. In addition, if the patterns of the 25^th^ or 75^th^ percentiles deviated from the median, the 90^th^ percentile was estimated. In a supplementary analysis, quartile-specific characteristics were computed to examine any potential effect of the intervention allocation or the different cohorts on the results.

### Ethical Approvals

The Trondheim Hip Fracture Trial was approved by the Regional Committee for Ethics in Medical Research in Central Norway (REK4.2008.335) and registered at ClinicalTrials.gov; NCT00667914. The Eva-Hip Trial was approved by the Regional Committee for Ethics in Medical Research in Central Norway (REK2010/3265-3) and registered at ClinicalTrials.gov; NCT 01379456. The PROFinD 1 trial had ethical approval from the University of Tübingen (113/2011BO2), and the PROFinD 2 trial from the Universities of Stuttgart (150/2015BO1) and Heidelberg (S-256/2015). All participants or proxies gave informed written consent to be included before participation in all trials. Reuse and merging of data from the Eva-Hip Trial was approved by the Regional Committee for Ethics in Medical Research in Central Norway after collecting passive consent from surviving participants (REK2022/ 412024). Data from the the Trondheim Hip Fracture Trial and the PROFinD 1 and PROFinD 2 trials were fully anonymised and available for reuse. All trials were conducted following the Declaration of Helsinki.

## Results

The current study included data from 717 hip fracture patients with a mean age of 83.4 (SD=6.1) years and 75.3% females, which was consistent across all cohorts. Average gait speed at four months (PROFinD 2 Trial: six months) post-surgery was 0.56 (SD=0.22) m/sec. Sample characteristics are shown in Table 1.

**Table 1.** Demographics and clinical characteristics of the patients included in the total sample and each chohort.

|  | Total |  | Trondheim Hip Fracture Trial | Eva-Hip Trial | PROFinD 1 Trial | PROFinD 2 Trial |
| --- | --- | --- | --- | --- | --- | --- |
|  | *n* = 717 | *n* | *n* = 357 | *n* = 130 | *n* = 93 | *n* = 137 |
| Age, years, mean (SD) | 83.4 (6.1) | 717 | 83.3 (6.0) | 83.2 (6.1) | 82.7 (6.5) | 84.4 (6.1) |
| Female gender, *n* (%) | 540 (75.3) | 717 | 269 (75.4) | 100 (76.9) | 68 (73.1) | 103 (75.2) |
| BMI, kg/m<sup>2</sup>, mean (SD) | 23.7 (4) | 427 | 23.4 (3.9) | 23.2 (3.9) | N/A | 24.6 (4.1) |
| Living alone at admission, *n* (%) | 443 (62.0) | 714 | 214 (59.9) | 95 (74.8) | 54 (58.1) | 80 (58.4) |
| Indoor falls, *n* (%) | 370 (72.5) | 510 | 213 (73.2) | 104 (82.5) | 53 (57.0) | N/A |
| Type of fracture, *n* (%) |  |  |  |  |  |  |
| FCF | 390 (54.4) | 717 | 219 (61.3) | 78 (60.0) | 41 (44.1) | 52 (38.0) |
| PTFF | 277 (38.6) | 717 | 113 (31.7) | 50 (38.5) | 43 (46.2) | 71 (51.8) |
| STFF | 50 (7.0) | 717 | 25 (7.0) | 2 (1.5) | 9 (9.7) | 14 (10.2) |
| Cognitive function* |  |  |  |  |  |  |
| MMSE (0-30), median (IQR) |  | 576 | 24 (8) | 26 (7) | N/A | 23 (4) |
| SOMC (0-28), median (IQR) |  | 93 | N/A | N/A | 2 (6) | N/A |
| Gait speed (preferred), m/sec, mean (SD)** | 0.56 (0.22) | 619 | 0.57 (0.22) | 0.63 (0.23) | 0.54 (0.23) | 0.49 (0.20) |

*Abbreviations:* SD, Standard Deviation; BMI, Body Mass Index; FCF, Fractura Collum Femoris; PTFF, Pertrochanteric Fractura Femoris; STFF, Subtrochanteric Fractura Femoris; Mini-Mental State Examination, MMSE (higher scores indicate better cognitive performance); Short Orientation Memory Concentration test, SOMC (lower scores indicate better cognitive performance); N/A, Not Available.
\*Four months post-surgery/ PROFinD 1 & 2 trials: three weeks post-surgery.
\*\*Based on the Short Physical Performance Battery's (SPPB) 4-meter walk at four (PROFinD 2 Trial: six) months post-surgery.

Throughout the year participants contributed with a total number of 5,909 observation days. The results of the quantile regression analyses for the three DMOs are presented in Figure 1 A-C.

**Figure 1.**
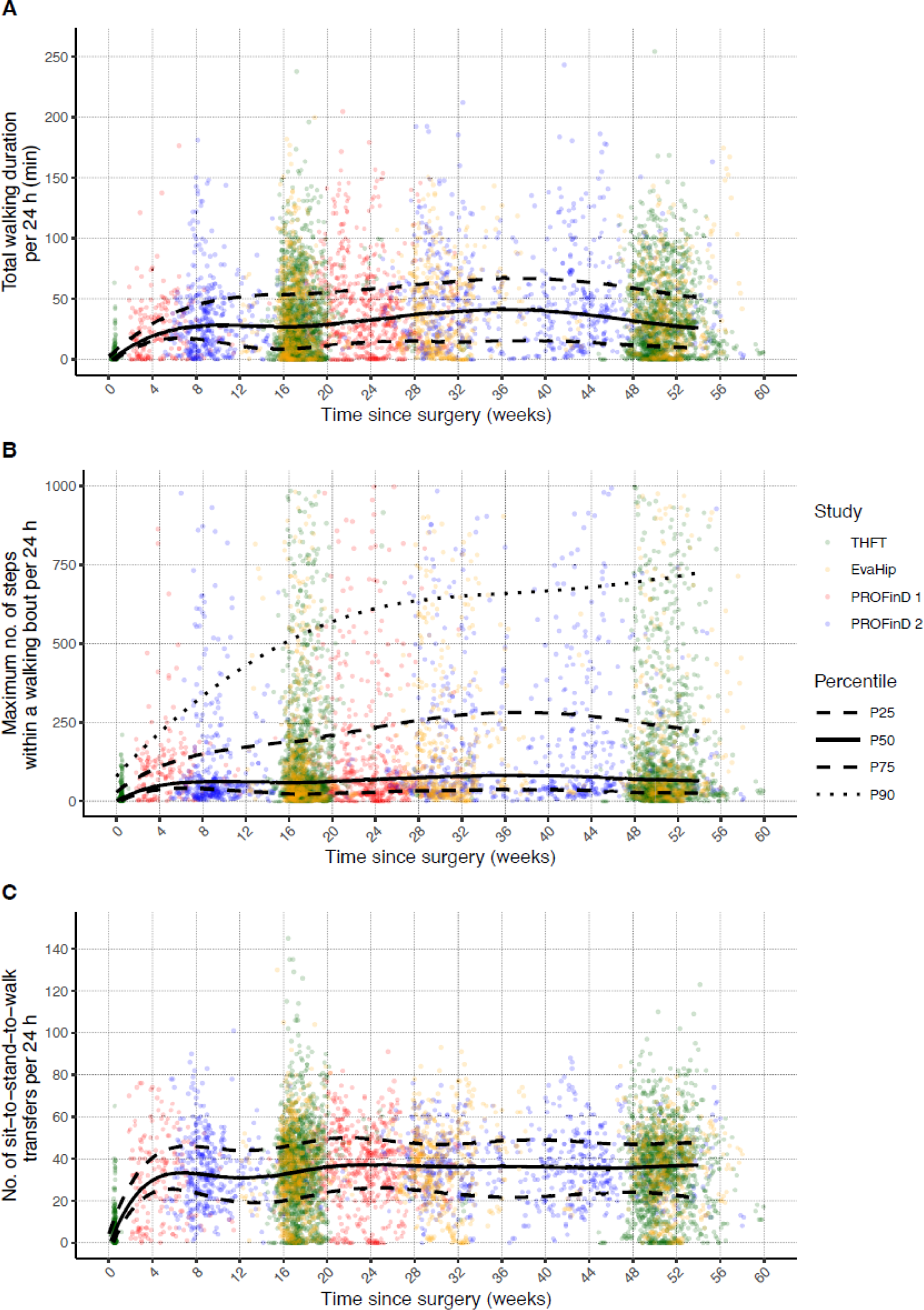
A-C. Quantile plots with the 25^th^, 50^th^ (median), and 75^th^ percentiles of the estimated one-year progression of the 24-hour DMOs total walking duration (A), maximum number of steps within a wallking bout (B), and number of sit-to-stand-to-walk transfers (C) for all four cohorts, and the 90^th^ percentile of the maximum number of steps within a walking bout (B). Green= Trondheim Hip Fracture lfrial (THFT); yellow= EvaHip Triial; red= PROFinD 1 lrial; blue= PROFinD 2 Trial.

All DMOs dispayed considerable variability. The 50^th^ percentile (median) of daily total walking duration increased until 36 weeks post-surgery, reaching 40 minutes (A). The most substantial increase occurred during the initial eight weeks post-surgery, followed by a decline starting at 36 weeks, eventually returning to the eight-week level, one year post-surgery. The 25^th^ and 75^th^ percentiles were mostly parallel to the median throughout. The 25^th^ percentile remained below 20 minutes.

The median of daily maximum number of steps within a walking bout (B) increased during the initial eight weeks and then stabilized at fewer than 100 continuous steps. The 75^th^ percentile increased until around week 36 before declining, while the 25^th^ percentile closely mirrored the median and did not exceed 50 steps. The 90^th^ percentile increased steadily throughout the year. Patients reached a plateau in their progress of daily transfers around six weeks post-surgery, with a median of less than 40. For this DMO the pattern of all quartiles was relatively parallel (C).

The supplementary data show the portion of measurement points within each quartile for each of the four included trials, stratified by gender and age (Appendix 1) and for the intervention groups only (Appendix 2), demonstrating few differences between trials. Visual inspection of the figure in Appendix 3 indicates an approximately equal distribution of measurement points from both the intervention and control groups across the quartiles throughout the year.

## Discussion

To our knowledge, this is the first study investigating one-year progression of real-world mobility recovery in patients after hip fracture using body-fixed movement sensors. By combining data from four previous trials involving 717 community-dwelling older hip fracture patients, we established a dataset with more than 5,900 observation days. Results of the estimated 24-hour total walking duration, maximum number of steps within a walking bout, and number of sit-to-stand-to-walk transfers showed that these three DMOs progressed differently and attained plateau levels at varying time points during the first year after hip fracture.

Over the course of one year, the DMOs showed different progression patterns of the median, emphasizing the necessity to consider assessment of various DMOs at different time points to gain insights into different aspects of real-world mobility recovery after hip fracture. For instance, the maximum number of steps within a walking bout reached a plateau within eight weeks, with fewer than 100 steps for half of the observations. This DMO may reflect an individual’s capacity for daily continuous steps and walking distance. Walking longer distances is vital for independence in especially outdoor activities, enabling patients to access necessary amenities. In contrast, accumulated daily walking duration increased as long as until week 36 post-surgery, with approximately 40 minutes or less for half of the observations. A daily walking duration of 40 minutes and a maximum of 100 continuous steps suggest that hip fracture patients do numerous shorter walks throughout the day. Additionally, the number of sit-to-stand-to-walk transfers rapidly reached a plateau after six weeks, with minimal change thereafter. These transfers play a critical role in upright mobility and are essential for maintaining functional independence. The median of approximately 40 transfers aligns with reports for frail older adults undergoing rehabilitation, but falls significantly below the almost 70 daily transfers observed in community-dwelling older adults [30], which could be considered a real-world rehabilitation target. Sit-to-stand-to-walk transfers may serve as a valuable proxy measure reflecting an individual’s ability to manage basic daily activities. The transfers appeared to stabilize once a certain threshold was reached, making them an appropriate outcome measure to assess both independence in upright mobility and decline in mobility impacting basic daily activities.

The maximum number of steps within a walking bout showed an increasing difference between the median and the 75th percentile until 36 weeks post-surgery. Interestingly, the upper ten percent of the observations continued to increase throughout the year. These findings underscore the heterogeneity of observations throughout the year, potentially reflecting patients’ differing rehabilitation needs and potential.

The 25^th^ percentile of total walking duration remained below 20 minutes, and the 25^th^ percentile of maximum number of steps within a walking bout did not exceed 50, followed by a decline around 6-8 weeks post-surgery. These observations suggest that the most vulnerable patients should be closely monitored. Given that most rehabilitation interventions typically end within the first four months post-surgery, there seems to be a need for evaluation of subsequent rehabilitation [31]. The considerable spread within our data indicates the need for personalised and targeted rehabilitation strategies to optimize patient care and enhance daily life mobility during hip fracture recovery. This aligns with a recent observational study that identified multiple patterns of mobility trajectories among older patients during in-patient hip fracture rehabilitation [32].

Due to the acute nature of hip fractures, real-world measures from body-worn sensors prior to the fracture are largely lacking. However, studies have shown that 40-60% of survivors fail to regain their self-reported mobility levels [5]. By monitoring mobility at different time points we were able to demonstrate low levels of real-world mobility throughout the entire year after hip fracture. Daily walking duration peaked at approximately 40 minutes, which is less than half of the average walking duration observed in a German study involving community-dwelling older adults with a mean age of 76 years [33], and is comparable to a study on frail and prefrail older adults with an average age of 81 years [34]. In contrast, an intervention study on hip fracture patients initiated approximately six months post-surgery found a significant increase in daily walking duration, reaching close to 65 minutes [17], emphasizing the importance of extended real-world activity monitoring beyond the initial six months. Monitoring real-world mobility helps identify changes, enabling personalized interventions and informing crucial windows for enhancing mobility in hip fracture patients.

A major strength of this study is the large sample with more than 700 individual hip fracture patients with a total of 5,909 observation days recorded with similar sensor systems. These observations were spread across the entire year after hip fracture, albeit with slight variations in distribution.

We acknowledge some methodological concerns. The activPAL algorithm has been shown to underestimate step counts and walking duration at slow gait speed [29, 35]. Given that the mean gait speed at 4-6 months post-surgery was 0.56 (SD 0.22) m/sec, this could have led to an underestimation of the walking outcomes for the slowest-walking participants.

We aggregated data from four intervention trials including both the intervention and control groups. The trials had minor differences in inclusion criteria, as well as different interventions at various intervals within the one-year follow-up period. These divergences could have influenced the results. However, Appendix 1 in the Supplementary data indicates that the observations from the various studies are distributed relatively equally within the quartiles, supporting the assumption that there were no cohort effects on the results.

Furthermore, based on visual inspection of the figure in Appendix 3 and information in Appendix 2 in Supplementary data, no significant effect of the interventions on quartiles is apparent. Finally, the clinical characteristics and demographic factors indicate a representative sample of hip fracture patients and thus a potentially high degree of external validity.

Our results are not based on trajectory estimations for individual patients. Observations from the same patient at different time points may potentially have contributed to different quartiles. However, at each point in time after hip fracture, the quartiles are the best estimates for the whole population. With continuous longitudinal data from each person across the whole observation period, DMO trajectories of similar subjects and their predictive factors could be explored in future studies.

## Conclusions and Implications

In summary, by monitoring real-world mobility we were able to gain insights into the patterns of recovery of mobility over the first year after hip fracture. The three DMOs progressed differently and attained plateau levels at varying time points, indicating that these DMOs provide complementary information about different aspects of mobility recovery and point to the necessity for various monitoring strategies at different time points.

The relatively limited activity observed throughout the year after hip fracture may point to a deficiency in targeted rehabilitation for certain individuals, possibly even for a majority of the patients. Hence, monitoring real-world mobility can provide deeper insight into mobility recovery after hip fracture, and provide valuable information to clinicians for tailoring rehabilitation to individuals.

## Data Availability

All data produced in the present study are available upon reasonable request to the authors.

## Declaration of Conflicts of Interest

The authors declare no conflicts of interest.

## Acknowledgments

The authors thank Olav Sletvold, Ingvild Saltvedt, and Pernille Thingstad who planned, led, and ran The Trondheim Hip Fracture Trial and The EVA-Hip Trial. We also thank Kilian Rapp, Klaus Pfeiffer, Karin Kampe, Michaela Kohler, Diana Albrecht, Klaus Hauer, Anja Dautel, Michaela Gross, Julia Gugenhan, Tobias Eckert, Bastian Abel, Andrej Lacroix for the collection and provision of the PROFinD 1 and 2 trials data sets.

## Notes

### Competing Interest Statement

The authors have declared no competing interest.

### Funding Statement

This work was funded by the Norwegian University of Science and Technology, Faculty of Medicine and Health Sciences, Trondheim, Norway, and the Mobilise-D project that has received funding from the Innovative Medicines Initiative 2 Joint Undertaking (JU) under grant agreement No. 820820. This JU receives support from the European Unions Horizon 2020 research and innovation program and the European Federation of Pharmaceutical Industries and Associations (EFPIA). Content in this publication reflects the authors view and neither IMI nor the European Union, EFPIA, or any Associated Partners are responsible for any use that may be made of the information contained herein. The Trondheim Hip Fracture Trial was funded by the Norwegian Research Council, the Central Norway Health Authority, the St Olav Hospital Trust, and the Department of Neuroscience at the Norwegian University of Science and Technology. The EVA-Hip Trial was funded by the Norwegian Womens Health Association and the Norwegian Extra Foundation for Health and Rehabilitation through the EXTRA funds, and the Norwegian Fund for Postgraduate Training in Physiotherapy. The PROFinD1 Trial was funded by the German Federal Ministry of Education and Research (grant number 01EC1007A). The PROFinD2 Trial was funded by the German Federal Ministry of Education and Research (grant number 01EC1404A-C).

### Author Declarations

The Regional Committee for Ethics in Medical Research in Central Norway gave ethical approval for this work (REK4.2008.335 and REK2010/3265-3). The ethical committees at the University of Stuttgart, and the Universities of Tubingen and Heidelberg gave ethical approval for this work (113/2011BO2, 150/2015BO1, and S-256/2015.

